# Genotype is a predictor of blood pressure variability and relative systemic hypertension risk in sickle cell disease

**DOI:** 10.64898/2026.06.06.26355049

**Authors:** Andre A.S. Bowers, Keneisha Henry, Brandon McConnell, Crystal Francis, Karen Thaxter-Nesbeth

## Abstract

**Background:** Blood pressure (BP) regulation in individuals with sickle cell disease (SCD) is influenced by a complex interplay of genetic and physiological factors. While SCD has traditionally been associated with lower BP, there is an increased risk of hypertension. Emerging BP research suggests significant heterogeneity across genotypes, age groups, and sex.

**Objectives:** This study investigated the longitudinal effects of population-level characteristics and continuous clinical and laboratory predictors on systolic (SBP) and diastolic blood pressure (DBP) in individuals with SCD, with emphasis on the interactions between baseline and predicted blood pressure slopes over time.

**Methods:** We retrospectively analyzed longitudinal data from a cohort of 2,739 patients with diverse SCD genotypes. Descriptive statistics were documented across sex, age range, genotype, health status and relative systemic hypertension risk categories (rHTN-risk). Linear mixed-effects models provided estimates of fixed- and random-effects of baseline BP and of time-related BP effects, respectively. Post-estimation margins provided contrasts of baseline-adjusted BP means and of pre-specified time effects on BP patterns.

**Results:** Males had significantly higher baseline SBP (β = 6.64, p < 0.001) but lower baseline DBP (β = -2.61, p < 0.001) compared with age-matched HbSS females. Baseline SBP was more unstable compared with baseline DBP and baseline DBP was more predictive of future BP trends than baseline SBP. Genotype was a consistent predictor of DBP (p < 0.05), but not of SBP. Similarly, we observed increased risks of relative diastolic hypertension across most genotypes, while the prevalence and magnitude of systolic hypertension was lower across all genotype compared with HbSS.

**Conclusions:** Blood pressure trajectories in SCD patients are not uniform and are significantly related to genotype, age group and sex over time. Baseline diastolic levels were less heterogenous and exhibited clear upward trajectories over time. These findings support the need for patient-specific BP surveillance in the care and management of SCD.

## INTRODUCTION

Sickle cell disease (SCD) describes a group of inherited blood disorders caused by the presence of abnormal haemoglobin S (HbS), which results from a single point mutation in the β-globin gene, substituting valine for glutamic acid at position six. The most severe and prevalent form, homozygous sickle cell disease (HbSS), manifests as chronic haemolytic anaemia, elevated circulating free haemoglobin, and nitric oxide depletion - mechanisms that promote endothelial dysfunction and recurrent vaso-occlusive crises [1–3]. Other clinically important genotypes include haemoglobin SC disease (HbSC), a compound heterozygote typically characterised by milder haemolytic anaemia but increased risks of proliferative retinopathy and thromboembolic complications. The HbS/β-thalassaemia syndromes arise from coinheritance of HbS with β-thalassaemia mutations [4].

In HbS/β^0^, there is complete absence of β-chain production, resulting in a clinical phenotype comparable to HbSS. HbS/β^+-^thalassaemia retains partial β-globin production, resulting in a milder phenotype, although complications such as vaso-occlusive pain and splenic dysfunction still occur [5]. Less common variants such as HbS/δ-β-thalassaemia or HbE/β-thalassaemia can produce a spectrum of clinical severity depending on the origin and extent of mutations [6].

Repeated cycles of erythrocyte sickling and vaso-occlusion, contribute to a persistent haemolytic state in SCD [4,7–10]. These processes markedly reduce erythrocyte and haemoglobin concentrations to levels that would be critical in the normal population [11–14]. However, unlike the non-SCD population where hypertension is prevalent, individuals with SCD are typically normotensive or even hypotensive compared to age-, race and sex-matched controls [15–19]. Nonetheless, a systolic blood pressure (SBP) ≥ 120 mmHg or a diastolic blood pressure (DBP) ≥ 70 mmHg in SCD is considered elevated and is a sign of increased relative hypertension (rHTN)-risk [20]. This paradoxical view of blood pressure (BP) distribution in SCD appear to demonstrate poorly understood genotype-dependencies, likely reflecting differences in disease severity. Individuals with clinically mild genotypes like HbSC and HbS/β^+^-thalassaemia, generally exhibit higher blood pressure compared with those with the more debilitating HbSS or HbS/β^0^-thalassaemia genotypes [20–21].

A 2021 cohort study of 442 adult SCD subjects and 884 NHANES control found that people with clinically milder genotypes - HbSC and HbS/β^+^- had higher median DBP compared with both HbSS subjects and HbAA controls [22]. These findings partly challenge the longstanding paradigm that patients with SCD, particularly HbSS, have lower median systolic and diastolic pressures compared with the non-SCD individuals [17,20,21,23]. Those authors attributed this historical trend to the chronic anaemia, reduced blood viscosity and the lower vascular tone, inherent to SCD pathophysiology.

Gordeuk et al. (2011), observed mean SBP of 103±10 mmHg and mean DBP of 65±8 mmHg in HbSS, compared with 109±11 mmHg and 68±7 mmHg in persons with the HbS/β^+^ genotype. These results are consistent with earlier findings and reaffirmed by a 2024 Nigerian paediatric study, which examined 106 children with HbSS and 106 age-matched controls (3 - 17 years) [24], further supporting the genotype-dependency of BP regulation in SCD. Disagreement between studies could be related to SCD-related heterogeneity across different cohorts. Interestingly, elevated blood pressure in individuals with HbSC has been linked with a disproportionately high prevalence of stroke and cerebral vasculopathy in females, but not males, based on findings in an 18-year retrospective study [25]. This is indicative of potential genotype-sex interaction in vascular risk that warrants further investigation.

Sex-related BP divergences in SCD appear to mirror patterns observed in the general pre-menopausal population [26–29]. However, in SCD, the direction and relative strength of these associations may vary by genotype and the presence of comorbid conditions. For instance, while body weight is positively associated with increased blood pressure in males, height does not appear to have an obvious effect. This lack of association may be related to delayed puberty and skeletal abnormalities [30] often observed in SCD or increased resting energy expenditure [31], which may potentially obscure any height-related vascular effects. Given these complexities, the interactions between genotype and sex in modulating blood pressure and hypertension-risk in SCD warrant further research. In particular, investigations into genotype- and sex-specific blood pressure interactions with common SCD comorbidities such as bone pain and nephropathy could provide useful insights into stratified hypertension-risk prediction in SCD.

We hypothesized that blood pressure in SCD is variably affected by genotype, sex, age and the presence of SCD-related comorbid complications. Therefore, we examined whether baseline blood pressure changed significantly over time based on these population-level covariates. Physiological predictors - weight, height, haemoglobin oxygen saturation and renal biomarker concentration - were included as time-varying continuous covariates. Regression models included random slopes and intercepts at the individual-level and random slopes for time. Subjects were categorised into low, high or severe relative hypertension-risk groups based on mean SBP and/or DBP values. Likewise, age was stratified into 10 year intervals, which enabled more generalized observation of BP distributions. Both Fixed- and random-effects (time) were included in the model as main covariates and interaction terms.

## MATERIALS & METHODS

### SUBJECTS AND DATA EXTRACTION

This was a retrospective longitudinal study of patient data documented and stored by the medical and clinical personnel, Sickle Cell Unit (SCU), University of the West Indies, Mona Campus, Jamaica, West Indies. A de-identified database containing blood pressure recordings collected between August 24, 2004 to August 24, 2013 was maintained at the SCU, Mona Campus, Kingston 7 by the clinic’s information technology personnel. Ethical approval for retrospective analyses was subsequently obtained in February 2026 from the University of the West Indies, Mona Campus, Research Ethics Committee, after which all statistical analyses were conducted. Strict measures for protection of identifying data and anonymity were employed throughout the process.

### HAEMOGLOBIN GENOTYPING

At the Sickle Cell Unit, Caribbean Institute of Health Research, University of the West Indies, Mona Campus, routine diagnostic tests comprised haemoglobin electrophoresis on citrate agar (pH 6.2) and cellulose acetate media, together with complete blood counts using automated analyzers (AC.Tron Coulter Counter).

### OUTCOME VARIABLES

We extracted information on continuous data including systolic and diastolic blood pressures, height, weight, haemoglobin oxygen saturation and biochemical markers - potassium (mmol/L), urea (mg/dL) and creatinine concentrations (mg/dL). Categorical variables comprised patients’ genotypes, sex and clinical history - including diagnoses of respiratory, renal, hepatic and cardiac complications, leg ulcers, infections and history of bone crises. The dataset was subsequently imported into Stata version 19 (Statacorp, Tx, USA) for data cleaning, variable coding and statistical analyses.

### DATA PREPARATION

The data consisted of systolic and diastolic blood pressure from 37,387 observations, each representing a separate patient visit. The dataset was inspected for coding inconsistencies, extreme outliers, normality-fit and duplicate entries tests using panel identifiers. Initially, a total of fifty-four observations were excluded from the dataset if; height (cm) ≥200, weight (kg) ≥150, systolic BP (mmHg) ≥200 or ≤70, diastolic BP (mmHg) ≥150 or ≤30. Another 3, 762 entries were excluded from the dataset due to duplication. A total of 33, 571 observations (55% females) remained - HbSS (76.4%); HbSC (15.47%), HbS/β^+^ (6.68 %) and HbS/β^0^ (1.45%).

Subsequently, 8, 169 observations were excluded if age < 18 years and 21 observations if age > 80 years. This yielded 25, 402 observations for baseline statistical analyses. Due to missing data on weight and oxygen saturation, multilevel models were restricted to 17, 762 observations. Data on height was not routinely documented in adults resulting in substantial missingness. Consequently, body mass index (BMI) was excluded from multilevel analyses. Height was analyzed separately where applicable.

Longitudinal models used visit number as the time index, which reflected clinical encounter-based progression in SCD care rather than actual calendar time. Each individual’s first recorded visit was treated as day zero, with successive measurements reflecting the number of days elapsed since this baseline. Because visit spacing was irregular and primarily associated with clinical needs, we performed sensitivity analyses replacing visit number with actual elapsed follow-up time (days since baseline). Results were consistent in direction and magnitude, supporting the robustness of our per-visit models. Logistic models used elapsed time to enable clinically interpretable per-time effects.

### RELATIVE HTN-RISK CATEGORIZATION

Mean age was stratified into six intervals: 18-29, 30-39, 40-49, 50-49, 60-69 and 70-79. Blood pressure was classified into three relative system hypertension-risk levels as either low-risk, high-risk or severe-risk based on the prior definitions of rHTN in SCD [20]: Low-risk: SBP < 120 & DBP < 70; High-risk: SBP ≥ 140 or DBP ≥ 90; Severe-risk: SBP ≥ 120 & SBP < 140 | DBP ≥ 70 & DBP < 90.

The steady state or routine visit was defined as a pre-scheduled visit for which there were no prevailing acute complications such as the acute chest syndrome or bone pain or crises. This did not account for 21 of 25,402 observations where routine scheduled visits may have coincided with presenting symptoms. Full details on demographic baseline characteristics, and BP stratification by sex, genotype and age group are provided in the Appendix.

### DATA ANALYSES

Summary table statistics consisted of number of observations (N), mean±SD for continuous variables (e.g. BP values) and frequency distribution with percentages (N (%)) for categorical variables (e.g. genotype and presence of comorbidity) (Figure 1; Appendix Table 1a). Covariates included sex, genotype, age group, height, weight, rHTN-risk categories, biochemical marker concentrations and SCD-related comorbidities. The two-sample t-test was used for pairwise comparisons. We used logistic regression models to test the time to event of binary outcomes according to rHTN-risk. Mean adjusted values were estimated in post-estimation analyses.

**Fig 1.**
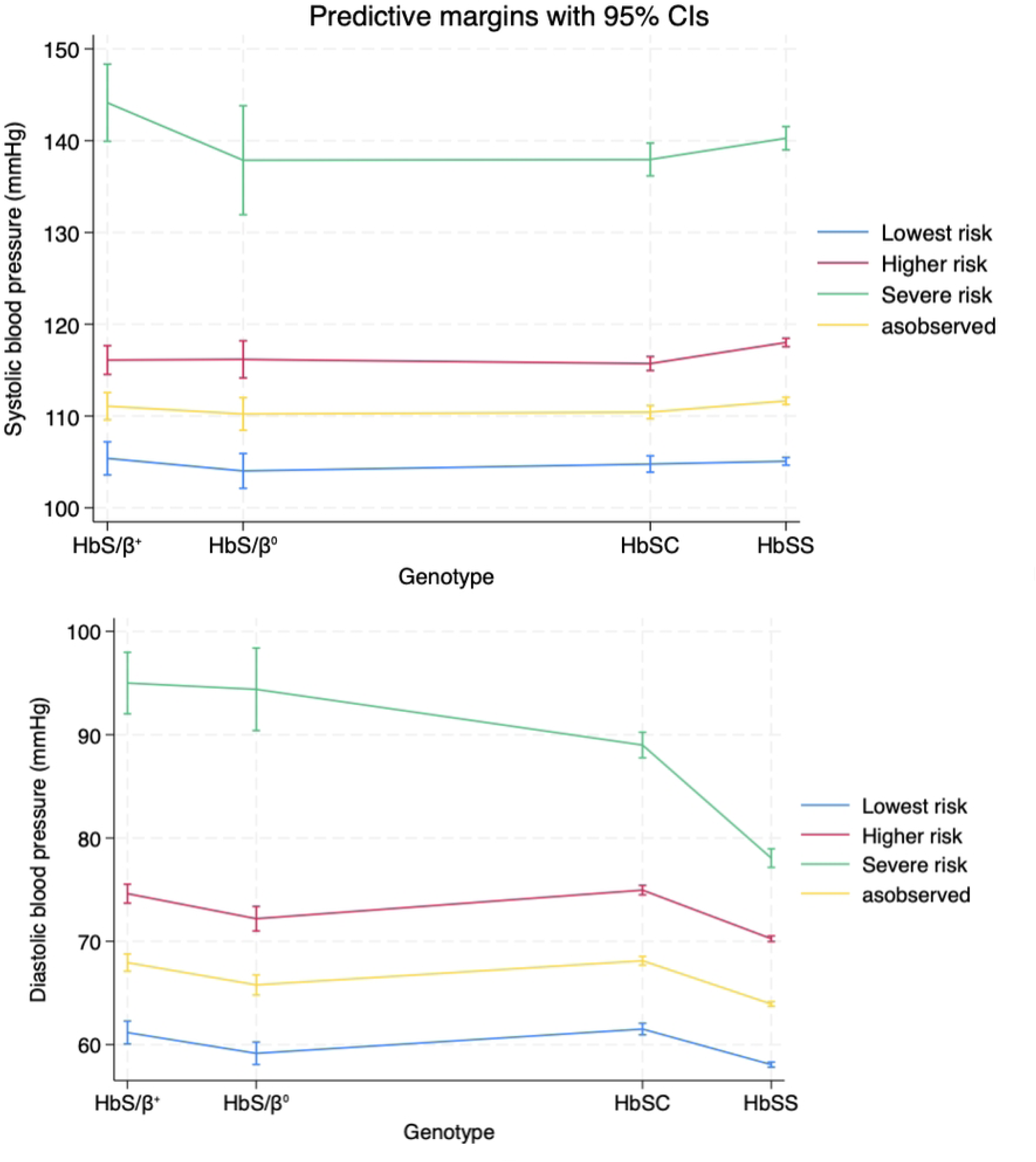
Mean-adjusted blood pressure (mmHg) by hypertension-risk strata and haemoglobin genotype.

**Table 1.**
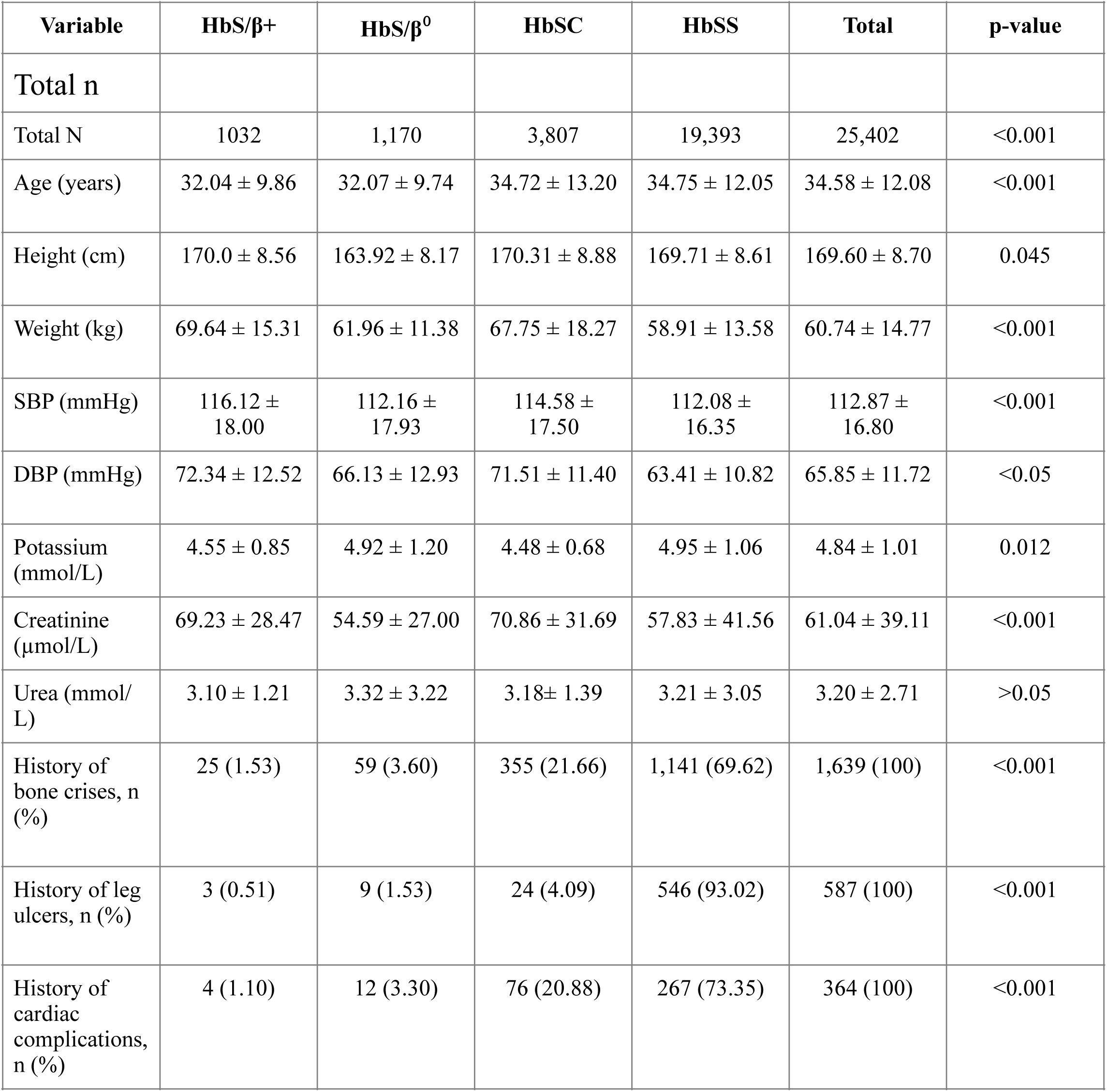
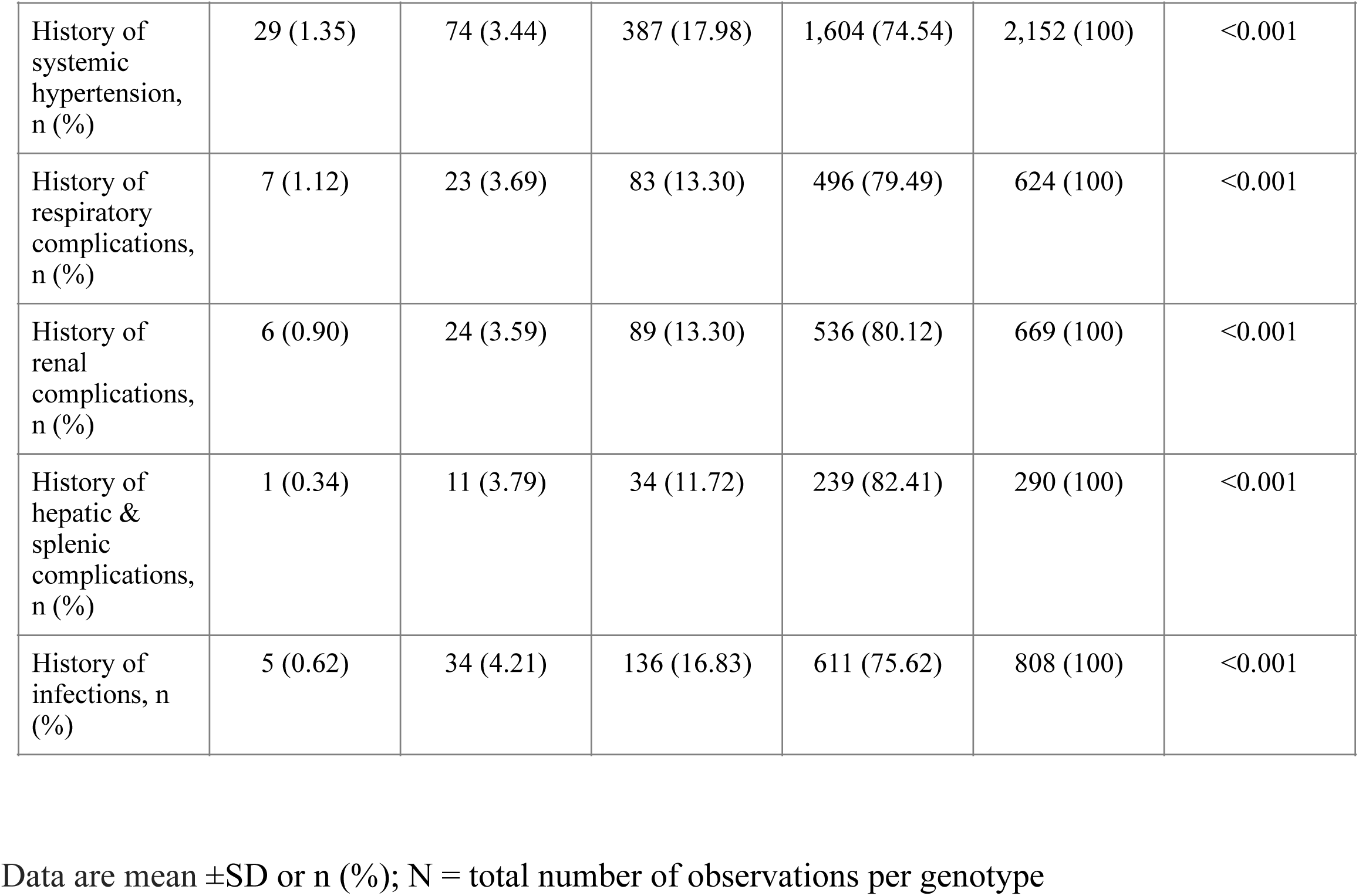
Summary of anthropometric, biochemical and clinical characteristics by haemoglobin genotype.

We evaluated the effects of genotype, sex, age and renal biomarker concentrations on blood pressure variability in SCD using linear mixed-effects regression models with an unstructured random-effects variance-covariance matrix. This approach was selected for its flexibility, as it imposes no parametric constraints on the variability or correlation structure across repeated measurements. Separate variances and covariances were estimated for random intercepts (baseline SBP/DBP) and random slopes (rates of change in SBP/DBP over time), allowing these components and their inter-relationships to be fully determined by the data. Regression interaction terms included genotype, rHTN-risk and time.

Fixed-effects included population-level variables (sex, genotype, rHTN-risk group and age) as well as time-varying covariates (weight (kg); potassium, urea and creatinine concentrations; presence or absence of comorbid conditions (renal, hepatic, cardiac, pulmonary, bone-pain related and leg ulcers)). Given the likelihood of some degree of autocorrelation across repeated measurements, we considered structures such as AR(1); however, uncertainty surrounding the correlation pattern for SCD blood pressure measurements favoured an unstructured covariance specification.

From these models, we obtained baseline-adjusted means and genotype-sex-time interaction combinations using post-estimation margins. We calculated pairwise differences with 95% CIs to quantify contrasts between genotypes and visit types (routine vs non-routine). Marginal plots provided visual illustration of mean-adjusted values across genotypes and hypertension strata. Where differences in scale could have confounded comparisons between systolic and diastolic fixed- and random-effects, coefficients of variation (CV) were additionally estimated to distinguish true heterogeneity from apparent differences due to scale.

To assess associations between blood pressure and clinically relevant binary outcomes, we fitted mixed-effects logistic regression with a patient level random intercept. Outcomes included rHTN-risk and comorbidity indicators in major organ systems, bone crises and infections. Models included the same fixed-effects described above, and where genotype and sex-specific associations were of interest, genotype-rHTN-risk and sex-rHTN interaction terms were incorporated. Odds ratios and 95% CIs were derived from model coefficients, and post-estimation margins were used to generate predicted probabilities. To examine whether the risk of rHTN and circulating renal biomarker levels were genotype-specific, we determined adjusted odds ratios with their 95% CIs.

All analyses were conducted using Stata (v.19.5, Statacorp, College Station, TX) with statistical significance set at p < 0.05 for two-sided tests and at the 1% level for odds ratios.

## RESULTS

### Study population

This study included data from 2,739 patients, comprising 25, 402 repeated clinic visits recorded between 2004 and 2013. Eligible persons had a confirmed diagnosis of homozygous sickle cell disease (HbSS), HbSC disease, HbS/β^0^-thalassaemia or HbS/β^+^-thalassaemia. Females accounted for 58.16% of all observations (p < 0.001) and were significantly older than presenting males on average (p < 0.001). HbSS was the most prevalent genotype, comprising 76.34% of total observations (p < 0.001), followed by HbSC disease at 15.0% (Table 1).

Overall median follow-up was 3.33 years (95% 3.27 5 to 3.38). Median follow-up was significantly shorter in individuals with BP values corresponding to high- (3.34 years; p = 0.041) and severe rHTN-risk (2.95 years; p = 0.002) compared to those with BP values corresponding to low-risk (3.35 years) over the 9-year study period. Furthermore, high-risk individuals had shorter follow-ups than the severe rHTN-risk group (p = 0.023).

Biochemical parameter concentration varied by genotype. Serum creatinine levels were significantly higher in HbSC compared with all other genotypes (p > 0.05). HbSS had significantly lower creatinine levels than HbS/β⁰ and HbSC (p < 0.05). Blood urea nitrogen concentrations were elevated in HbSC compared with HbS/β^+^, and HbSS (p < 0.05). Potassium levels were comparable across genotypes (p > 0.05).

The cumulative prevalence of chronic comorbidities was 74.37% (N = 2,036; 95% CI: 72.7–76.0%): leg ulcers (N=1639, 59.8%), infections (N=808, 29.5%), systemic hypertension (N=220, 8%), renal dysfunction (N=669, 24.4%), liver and/or spleen complications (N=290; 10.6%) and cardiac involvement (N=364, 13.3%) (Table 1).

### Genotype, sex and age effects

Mean blood pressure varied according to haemoglobin genotype as follows - HbS/β⁺ > HbSC > HbS/β^0^ > HbSS (Table 1). In multivariate analyses, haemoglobin genotype was not an independent predictor of SBP, whereas DBP was significantly greater in HbS/β^0^ (β = 1.18; p = 0.047), HbS/β^+^ (β = ; p = 0.0) and HbSC (β = 3.33; p = 0.001) compared with HbSS. On average, SBP was 2.65 mmHg higher in males than females (β = 2.65, p < 0.001), whereas mean DBP was higher in females than males (β = -0.91, p < 0.001) regardless of genotype (Table 2).

**Table 2:** Mixed effects analyses of the effects of genotype, sex and visit number and context on systolic pressure in SCD.

| Predictor | SBP Coefficient (p-value) | DBP Coefficient (p-value) |
| --- | --- | --- |
| Genotype-associated change: |  |  |
| HbS/$\beta^+$ | 0.69 (p = 0.50) | 3.11 (p < 0.001) |
| HbS/$\beta^0$ | -1.36 (p = 0.184) | 1.19 (p = 0.047) |
| HbSC | -0.39 (p = 0.470) | 3.33 (p < 0.001) |
| High rHTN-risk versus HbSS | 13.32 (p < 0.001) | 11.83 (p < 0.001) |
| Severe rHTN-risk versus HbSS | 36.38 (p < 0.001) | 19.35 (p < 0.001) |
| Sex-related change: Male | 2.63 (p < 0.001) | -0.91 (p < 0.001) |
| Visit number-related change | -0.01 (p = 0.291) | 0.01 (p = 0.182) |
| Visit type-related change | -0.02 (p = 0.95) | -0.13 (p = 0.507) |
| Weight associated change | 0.06 (p < 0.001) | 0.04 (p < 0.001) |
| Age-related change: |  |  |
| Age 30–39 | 0.93 (p = 0.005) | 1.42 (p < 0.001) |
| Age 40–49 | 3.69 (p < 0.001) | 1.78 (p < 0.001) |
| Age 50–59 | 9.44 (p < 0.001) | 1.51 (p < 0.001) |
| Age 60–69 | 16.24 (p < 0.001) | 1.55 (p = 0.014) |
| Age 70–79 | 15.50 (p < 0.001) | -1.80 (p = 0.135) |

\*Genotype was included as the random effect and the model had main effects only.

Age was a consistent predictor predictor of both SBP and DBP (p < 0.05). While differences between age groups were small in magnitude, SBP showed substantial progressive increases across age strata. Systolic blood pressure values corresponding to low- and high-risk hypertension categorization were, on average, lower in HbSS by 13.32 mmHg and 36.38 mmHg, respectively (p < 0.001).

#### Mean-adjusted blood pressure by visit type

After adjusting for sex, age and other covariates, mean-adjusted systolic pressure was significantly elevated during non-routine visits in persons with HbSS (MD -1.09; 95% CI -1.75 to -0.43). In contrast, adjusted SBP was generally higher during non-routine visits, although this difference was not statistically significant ([MD] range: -1.15 to -0.20; 95% CI -3.70 to 2.85) (Table 3; Figure 1). Diastolic pressure also tended to be higher in persons visiting for acute events compared with routine visits ([MD] range: -0.65 to 0.04; 95% CI -2.16 to 1.27).

**Table 3.** Baseline-adjusted blood pressure by genotype and visit type.

| Genotype | Routine<br>(mean) | Routine<br>(95% CI) | Non-routine<br>(mean) | Non<br>routine CI<br>(95% CI) | Mean<br>difference | 95% CI |
| --- | --- | --- | --- | --- | --- | --- |
| **Systolic** | **Blood** | **Pressure** |  |  |  |  |
| HbS/$\beta^+$ | 110.21 | 108.21 to<br>112.21 | 111.36 | 109.79 to<br>112.95 | -1.15 | -3.70 to<br>1.40 |
| HbS/$\beta^0$ | 110.50 | 108.02 to<br>112.99 | 110.70 | 108.93 to<br>112.47 | -0.20 | -3.25 to<br>2.85 |
| HbSC | 109.92 | 108.94 to<br>110.90 | 110.55 | 109.78 to<br>111.32 | -0.63 | -1.88 to<br>0.62 |
| HbSS | 110.75 | 110.23 to<br>111.28 | 111.84 | 111.44 to<br>112.25 | -1.09 | -1.75 to<br>-0.43 |
| **Diastolic** | **Blood** | **Pressure** |  |  |  |  |
| HbS/$\beta^+$ | 67.70 | 66.44 to<br>68.98 | 68.00 | 67.08 to<br>68.93 | -0.3 | -1.87 to<br>1.27 |
| HbS/$\beta^0$ | 65.30 | 63.69 to<br>66.91 | 65.95 | 64.96 to<br>66.95 | -0.65 | -2.16 to<br>0.86 |
| HbSC | 68.15 | 67.52 to<br>68.78 | 68.11 | 67.65 to<br>68.57 | 0.04 | -0.73 to<br>0.81 |
| HbSS | 63.87 | 63.53 to<br>64.21 | 63.93 | 63.70 to<br>64.17 | -0.06 | -0.39 to<br>0.27 |

N=number of visits per group, n=number of participants; values are mean ± sd, median(iqr); MD: routine - non-routine; positive values indicate higher BP during routine visits. N=number of visits per group, n=number of participants; values are mean ± sd, median(iqr); MD: routine - non-routine; positive values indicate higher BP during routine visits.

#### Hypertension-risk stratification

Genotype emerged as a significant independent predictor of relative systolic hypertension in post-regression analyses. Compared with HbSS, individuals with HbS/β^+^ (β=-2.23; 95% CI -3.91 to -0.53; p = 0.010 and β=3.55; 95% CI -0.91 to 8.01; p = 0.119), HbS/β^0^ (β=-0.79; 95% CI -2.41 to 0.83; p = 0.340 and β=-1.35; 95% CI -7.41 to 4.71; p = 0.662) and HbSC (β=-2.00; 95% CI -2.90 to -1.10; p < 0.001 and β=-2.02; 95% CI -4.24 to 0.20; p = 0.075), had significantly lower SBP values corresponding to lower risks of relative systemic hypertension in both the high- and severe-risk categories, respectively. Only HbS/β^+^ had SBP values corresponding to lower severe rHTN-risk compared with HbSS.

Post-estimation analyses revealed pronounced genotype-specific associations in relative diastolic hypertension-risk, consistent with linear model predictions - HbS/β^+^ (β=1.26; 95% CI 0.07 to 2.44; p = 0.037 and β=13.82; 95% CI 10.63 to 17.04; p < 0.001) and HbS/β^0^ (β=0.84; 95% CI -0.32 to 2.01; p = 0.155 and β=15.24; 95% CI 11.10 to 19.38; p < 0.001) and HbSC (β=1.26; 95% CI 0.62 to 1.89; p < 0.001 and β=7.49; 95% CI 5.92 to 9.07; p < 0.001) were strongly associated with both high and severe rHTN-risk, respectively, compared with HbSS. (Table 4) (Figure 1). Individuals with HbS/β^0^ were associated with a low probability of high-risk rHTN.

**Table 4.** Adjusted blood pressure means by genotype and relative systemic hypertension-risk.

| **Genotype** | **Lowest Risk (95% CI)** | **Higher Risk (95% CI)** | **Severe Risk (95% CI)** | **Δ Higher - Lowest (95% CI)** | **Δ Severe - Lowest (95% CI)** |
| --- | --- | --- | --- | --- | --- |
| **Systolic** | **blood pressure** |  |  |  |  |
| HbS/β<sup>+</sup> | 103.65 (102.99 to 107.39) | 116.24 (114.64 to 117.84) | 144.13 (139.93 to 148.33) | 12.59 (7.25 to 14.85) | 40.48 (32.54 to 45.34) |
| HbS/β<sup>o</sup> | 104.04 (102.14 to 105.94) | 116.43 (114.33 to 118.53) | 137.87 (131.93 to 143.80) | 12.39 (8.39 to 16.39) | 33.83 (25.99 to 41.66) |
| HbSC | 104.74 (103.84 to 105.63) | 115.76 (114.99 to 116.53) | 138.13 (136.19 to 140.08) | 11.02 (9.36 to 12.69) | 33.39 (30.56 to 36.24) |
| HbSS | 105.05 (104.63 to 105.47) | 117.95 (117.50 to 118.40) | 139.50 (138.18 to 140.82) | 12.90 (12.03 to 13.77) | 34.45 (32.71 to 36.19) |
| **Diastolic** | **blood pressure** |  |  |  |  |
| HbS/β<sup>+</sup> | 60.97 (59.83 to 62.11) | 74.60 (73.67 to 75.54) | 94.99 (92.02 to 97.97) | 13.63 (11.56 to 15.71) | 34.02 (29.91 to 38.14) |
| HbS/β<sup>o</sup> | 59.20 (58.12 to 60.29) | 72.27 (71.01 to 73.52) | 94.39 (90.40 to 98.39) | 13.07 (10.72 to 15.40) | 35.19 (30.11 to 40.27) |
| HbSC | 61.46 (60.90 to 62.02) | 74.94 (74.49 to 75.40) | 89.41 (88.06 to 90.77) | 13.48 (12.47 to 14.50) | 27.95 (26.04 to 29.87) |
| HbSS | 58.08 (57.83 to 58.32) | 70.28 (70.01 to 70.56) | 77.81 (76.88 to 78.74) | 12.20 (11.69 to 12.73) | 19.73 (18.56 to 20.91) |

Values represent mean(95% CI) and hypertension-risk and mean risk difference from baseline values across SCD genotypes.

#### Blood pressure variability over time

Although baseline variability was higher for SBP (SD = 6.81 mmHg; 95% CI 6.48 to 7.17) than DBP (SD = 3.52 mmHg; 95% CI 3.31 to 3.75), coefficients of variation (CV) were comparable (5.9% versus 5.0%), suggesting that the apparent disparity largely reflected scale rather than disproportionate dispersion. In contrast, inter-individual variability in visit-related slopes was nearly two-fold greater for SBP (SD = 0.154 mmHg/visit; 95% CI 0.112 to 0.211) than for DBP (SD = 0.079 mmHg/visit; 95% CI 0.054 to 0.116), indicating genuine heterogeneity and greater instability in SBP patterns over time.

Whereas cross-sectional intercepts were weakly and non-significantly correlated with SBP slopes (r = -0.17; 95% CI -0.37 to 0.04), DBP intercepts were significantly and negatively correlated with DBP slopes (r = -0.35; 95% CI -0.53 to -0.13). This meant that individuals with higher baseline DBPs tended to experience steeper DBP reductions over time. Within-person residual variability - representing unexplained visit-to-visit fluctuations not accounted for by the model - was also higher for SBP (SD = 8.85 mmHg; 95% CI 8.75 to 8.95) than for DBP (SD = 6.45 mmHg; 95% CI 6.38 to 6.53).

Fixed-effects estimates from the SBP model showed no significant genotype-time or sex-time interactions. However, individuals with severe rHTN-risk had significantly higher SBP than the reference group by 36.7 mmHg (95% CI 35.41 to 38.02; p < 0.001). Males had an average per-visit SBP of 2.64 mmHg higher than age- matched females (95% CI 1.94 to 3.33). Conversely, DBP was 1.16 mmHg lower in males over the same period (95% CI -1.58 to -0.74; p < 0.001).

Over time, both high (β=-0.04; (95% CI -0.06 to -0.03; p < 0.001)) and severe (β=-0.15; (95% CI -0.21 to -0.09; p < 0.001)) diastolic rHTN-risk categories were associated with significant downward trends, consistent with the observed negative intercept-slope correlation pattern in the DBP models (Figure 2). Notably, individuals with HbS/β^+^ had a significant 0.30 mmHg increase in diastolic pressure between visits compared with HbSS.

**Fig 2.**
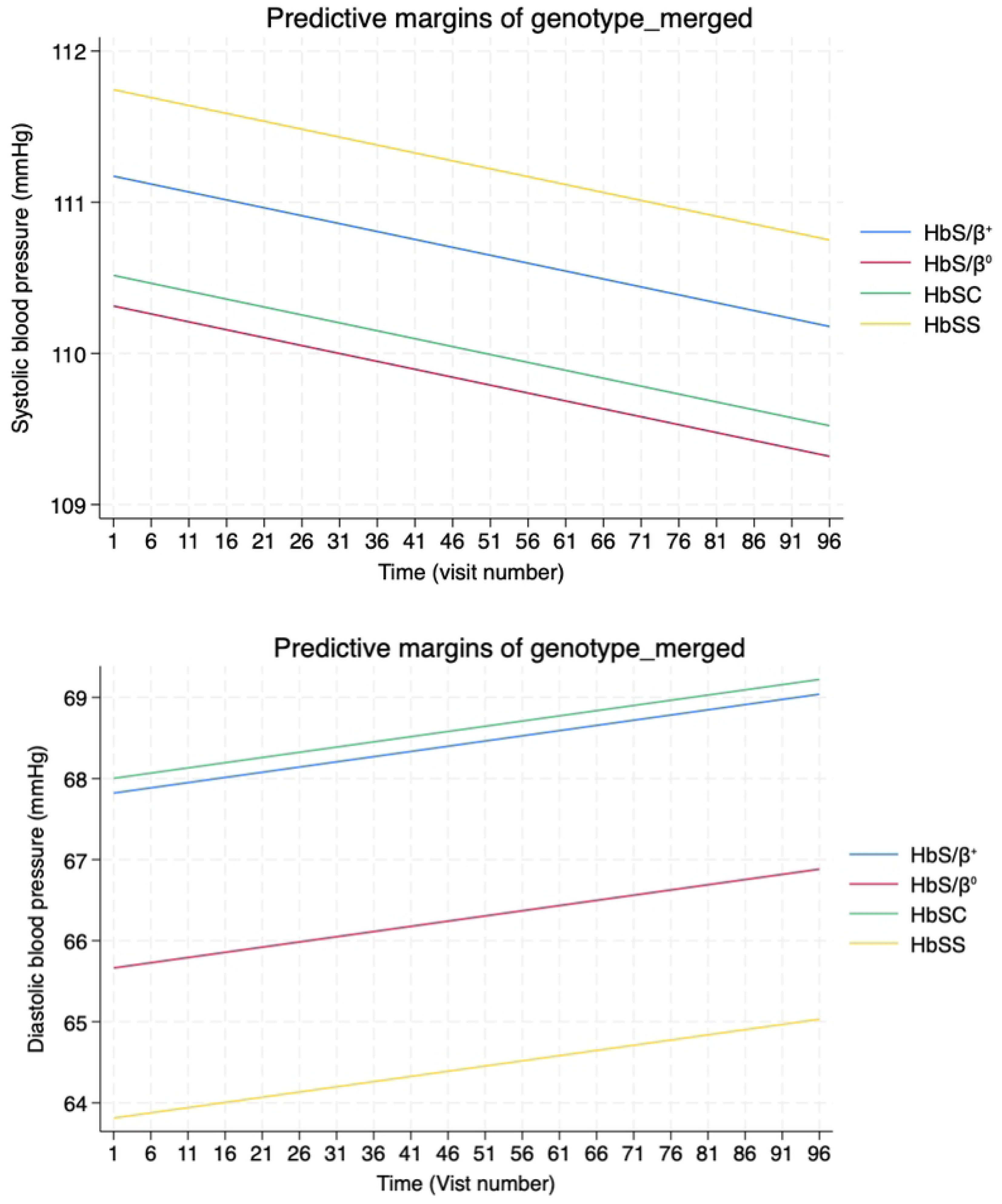
Genotype-associated mean-adjusted blood pressure over time in sickle cell disease. Post-estimation predictive margins of SBP and DBP across SCD genotypes. Values represent baseline estimates to the 500th visit. While DBP margins increased with repeated visits and showed clear genotype divergences, SBP margins decreased over the same period and showed less genotype-specific variability.

### Genotype-specific biochemical interactions

Genotype-specific biochemical interactions were largely negatively associated with predicted systolic blood pressure, suggesting higher circulating concentrations of certain biomarkers generally corresponded to higher SBP in HbSS (Table 5). In multivariate analysis, potassium (MD -1.218 (95% CI -2.392 to -0.045) and creatinine (MD -0.033 (95% CI -0.066 to -0.001) concentrations in HbSC and urea in HbS/β^0^ (MD -0.377 (95% CI -0.755 to 0.002) were significantly lower than the respective values observed in HbSS. Conversely, creatinine was significantly elevated in HbS/β^0^ (MD 0.061 (95% CI 0.011 to 0.111) relative to HbSS.

**Table 5.** Estimated interaction effects between genotype and biochemical markers on systolic and diastolic blood pressure.

| **Genotype × Biomarker** | **SBP Estimate (95% CI)** | **SBP p-value** | **DBP Estimate (95% CI)** | **DBP p-value** |
| --- | --- | --- | --- | --- |
| HbS/β<sup>+</sup> × Potassium | 0.53 (-1.47 to 2.53) | 0.604 | 0.01 (-6.41 to 6.43) | 0.997 |
| HbS/β<sup>0</sup> × Potassium | -0.28 (-2.280 to 1.720) | 0.783 | 1.804 (0.428 to 3.180) | 0.01 |
| HbSC × Potassium | -1.218 (-2.392 to -0.045) | 0.042 | -0.79 (-1.598 to 0.018) | 0.055 |
| HbS/β<sup>+</sup> × Creatinine | 0.04 (-0.02 to 0.09) | 0.172 | 1.12 (-0.25 to 2.50) | 0.110 |
| HbS/β<sup>0</sup> × Creatinine | 0.061 (0.011 to 0.111) | 0.016 | 0.06 (0.026 to 0.094) | 0.001 |
| HbSC × Creatinine | -0.033 (-0.066 to -0.001) | 0.043 | -0.004 (-0.026 to 0.019) | 0.75 |
| HbS/β<sup>+</sup> × Urea | -0.78 (-1.73 to 0.17) | 0.106 | 0.08 (0.04 to 0.12) | 0.001 |
| HbS/β<sup>0</sup> × Urea | -0.377 (-0.755 to 0.002) | 0.051 | -0.425 (-0.685 to -0.165) | 0.001 |
| HbSC × Urea | 0.116 (-0.557 to 0.789) | 0.735 | 0.044 (-0.419 to 0.507) | 0.852 |

Values represent genotype-biomarker interaction coefficients $\beta$ (95% CI) and p-values of systolic and diastolic pressures mean across SCD genotypes.

Associations with diastolic pressure predominantly showed positive trends, indicating that lower biomarker concentrations were characteristics of HbSS. Specifically, potassium (MD 1.804 (95% CI 0.428 to 3.180) and creatinine (MD 0.06 (95% CI 0.026 to 0.094) in HbS/β^0^ and urea (MD 0.08 (95% CI 0.04 to 0.12) in HbS/β^+^ were significantly elevated compared with corresponding values in HbSS. In contrast, potassium in HbSC (MD -0.79 (95% CI -1.598 to 0.018) and urea in HbS/β^0^ (MD -0.425 (95% CI -0.685 to -0.165) were significantly reduced compared with HbSS.

Briefly, a one unit increase in potassium concentration was associated with a 4.72 mmHg rise in SBP in HbS/β^+^ compared with HbSS (p = 0.022), whereas HbSC showed a modest -1.42 mmHg decrease (p = 0.042; Table 4). Creatinine levels were positively associated with SBP in HbS/β^0^ (β=0.06; p = 0.016). While SBP appeared more sensitive to potassium and creatinine changes, DBP was more susceptible to variation in urea and creatinine concentrations, particularly among the HbS/thalassaemia genotypes. Among persons with HbSC, creatinine was linked to significantly lower DBP ( β=-0.033, p = 0.043; β=1.22, p = 0.042), while urea showed no meaningful associations (Table 5).

#### Comorbidity-specific predictors of hypertension-risk

The distribution of comorbid complications varied markedly across rHTN-risk groups. In the low and high rHTN-risk categories, bone pain, followed by leg ulcers, and infections accounted for majority of recorded complications. In the severe rHTN-risk group - bone crises > leg ulcers > cardiac disease > kidney involvement - were most frequently observed (Figure 3).

**Fig 3.**
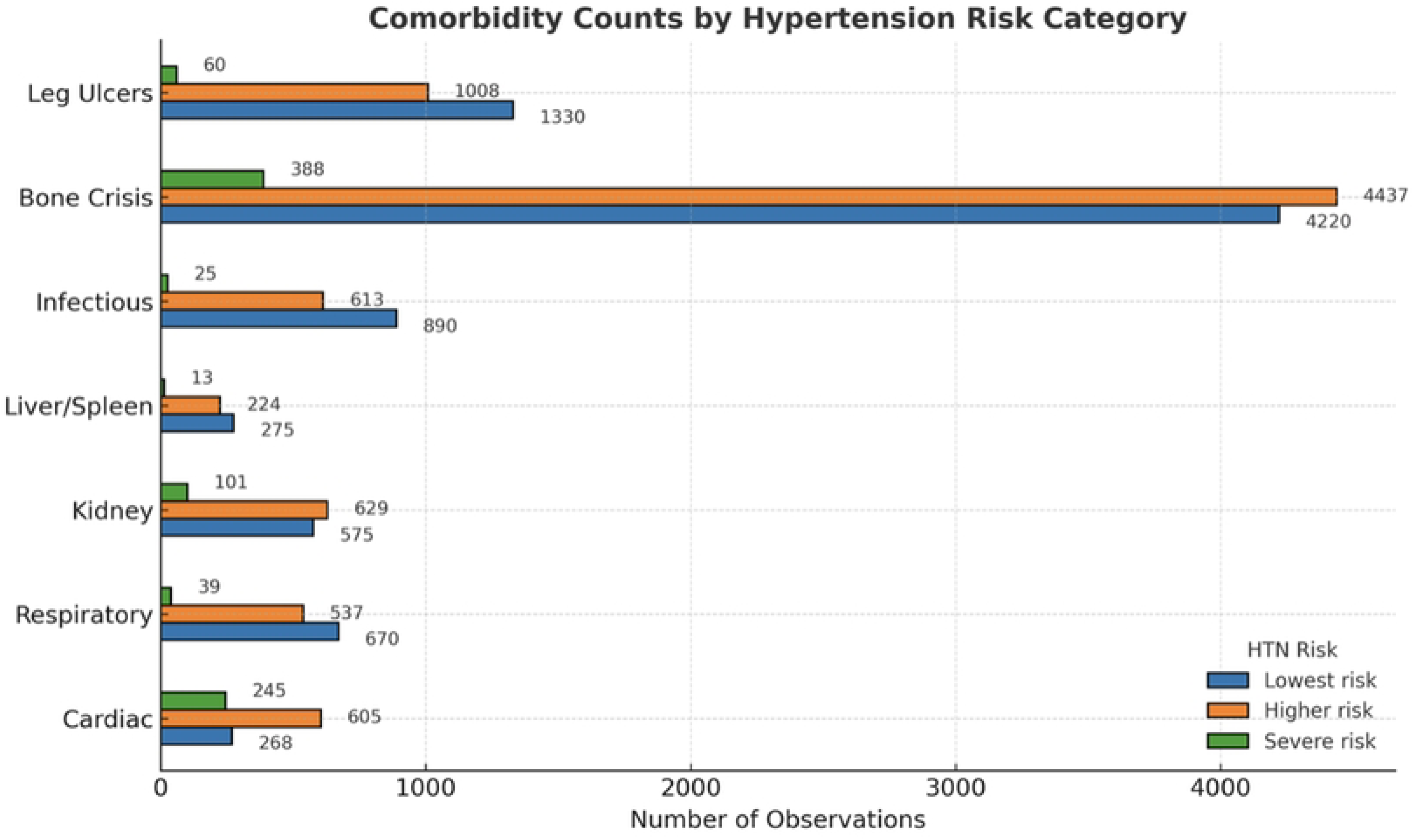
Frequency of SCD-related comorbidities by hypertension-risk categories. Bars represent number of observations per comorbidity and hypertension-risk class. Total observations: lowest risk - 13,161; higher risk - 11,405; severe risk - 836.

Logistics regression showed that cardiac complications were associated with a 49% increase in the odds of hypertension (OR = 1.49; 95% CI 1.31 to 1.68; p = 0.001), 10% higher than the odds conferred by bone crises (OR = 1.39; 95% CI 1.31 to 1.46; p = 0.001) and more than double the predictive effect of kidney disease relative to the low-risk group (OR = 1.22; 95% CI 1.09 to 1.37; p = 0.001). Respiratory and hepatic disease were unrelated to hypertension-risk, while infections appear to have had a protective effect (OR = 0.85; 95% CI 0.76 to 0.95; p = 0.003).

In the severe risk category, kidney disease (OR = 2.04; 95% CI 1.61 to 2.58; p = 0.001) and bone crises (OR = 2.20; 95% CI 1.89 to 2.57; p = 0.001) doubled the odds of hypertension, while cardiac disease was associated with a striking 12-fold increase in hypertension-risk (OR = 12.09; 95% CI 10.14 to 14.42; p = 0.001). Hepatic and respiratory dysfunction remained non-significant, while infections retained a protective effect, lowering the odds of rHTN by 39% (OR = 0.61; 95% CI 0.41 to 0.92; p = 0.018) (Figure 4).

**Fig 4.**
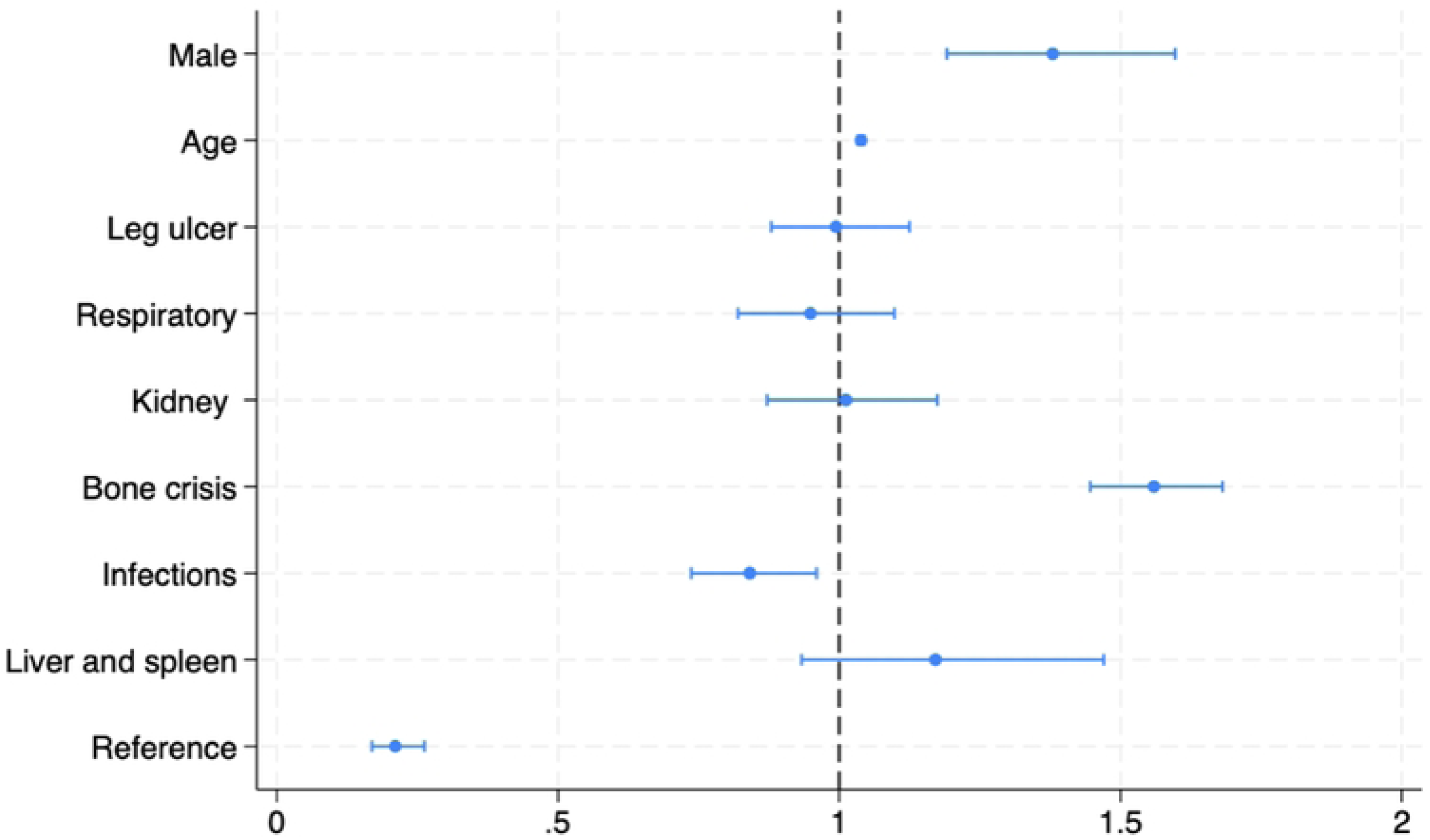
The overall odds of relative systemic hypertension in sickle cell disease. Indicators to the left of the no effect line (> 1) represent reduced odds; those > 1 represent increased odds. Values are significant if 95% CIs exclude 1.

The odds of systemic hypertension varied according to haemoglobin phenotype as follows: HbS/β^+^ (OR 6.31; 2.94 to 13.55; p < 0.001) > HbSC (OR 3.91; 3.10 to 4.93; p < 0.001) > HbS/β^0^ (OR 1.50; p = 0.106) > HbSS (OR ; p = 0.00).

## DISCUSSION

To our knowledge, this study provides the most detailed genotype- and phenotype-specific characterisation of blood pressure distribution in sickle cell disease to date, with particular attention on non-haematological indicators of disease severity. It is also the first to link time-related variability, and the observed divergences between systolic and diastolic pressure, to common clinical outcomes and established indicators of increased morbidity - including a history of bone crises, cardiac complications and nephropathy - as well as to circulating biomarkers suggestive of renal dysfunction. Situational differences in blood pressure (routine versus non-routine) were generally not significant, except a significant 1.09 mmHg difference in HbSS. A documented history of certain comorbidities markedly increased the risk of relative systemic hypertension (rHTN), with prevalence patterns varying systematically across low-, high- and severe-risk categories. Cardiac complications were associated with a 49% increase in the odds of high-risk rHTN and a striking 12-fold increase in the odds of severe rHTN, while bone crises and kidney disease both conferred a two-fold increase in the odds of severe rHTN.

### Blood pressure dysregulation in SCD

The pathophysiologic basis of blood pressure (BP) dysregulation in SCD involves a complex interplay of factors, including autonomic dysfunction, cardiovascular abnormalities [16,20,27,29], nephropathy [32–34], increased vascular tone [35] and baroreceptor insensitivity [36]. Peripheral vasoconstriction plays a mitigating role in maintaining adequate blood pressure. However, in the absence of adequate cardiac output, due to autonomic impairment and chronic haemolysis, sickling and vaso-occlusion ensues [37–38]. These inter-related mechanisms contribute to lower baseline blood pressure than published standards, but with a state of heightened organ sensitivity, whereby even mild perturbations in blood pressure or haemolytic rate can result in disproportionate end-organ damage [20,39–40].

### The effects of genotype, age and sex

After adjusting for covariates, systolic blood pressure rose progressively with age and was higher in males, but showed little sensitivity to genotype or longitudinal change. In contrast, diastolic blood pressure was more genotype-dependent, lower in males and demonstrated significantly greater modification over time and with clinical state (e.g. HTN-risk level). Although mean BP values during acute events such as bone pain crises were marginally higher - particularly for systolic pressure - and reached statistical significance in ordinary least squares regression, these differences were attenuated and did not remain statistically significant in multivariate models.

The physiological basis of higher SBP during painful vaso-occlusive episodes likely involves a sympathetic surge, with catecholamine release driving transient elevations in systolic pressure [41–43]. In SCD, however, these responses are blunted by vasodilatory impairment related to chronic oxidative stress [44–45], reduced NO-bioavailability and endothelial dysfunction [46–49]. Chronic anaemia, a proximate driver of this oxidant state, promotes vasoconstriction and a pro-inflammatory endothelial phenotype [49–50], contributors to vascular dysfunction and large-artery stiffening - effects that are most pronounced in older patients. Additional contributors include dehydration and hypovolemia, often arising from tubular hyperfiltration and nephropathy [51]. Renal abnormalities exacerbate BP dysregulation and may intensify pain perception through vasoconstriction mediated by the activation of the renin-angiotensin-aldosterone-system (RAAS) [19,52]. Finally, disruption of ACE-mediated angiotensin II conversion could further influence pain pathways, adding another layer of complexity to BP dysregulation during vaso-occlusive crises.

These mechanisms help explain the differential trajectories of SBP and DBP in this cohort; SBP, dominated by stroke volume and large-artery load, remained stable longitudinally but displayed context-related surges consistent with increased sympathetic activity and fluid shifts during acute care. DBP, by contrast, more closely reflected changes in peripheral resistance and microvascular tone. Its stronger associations with genotype and clinical risk factors suggest that DBP, rather than SBP, may serve as a predictor of evolving vasculopathy in SCD, marking a departure from conventional hypertension paradigms.

Sex differences further refined these BP patterns: Higher SBP in males was likely related to higher stroke volume due to larger body mass [21,53,54]; Lower DBP in males suggest lower vascular tone, possibly mediated by increased β-adrenergic vasodilation [55]. These haemodynamic patterns align with the high-output state of chronic anaemia [21], wherein lower haematocrit reduces whole blood viscosity, which lowers vascular resistance, thereby facilitating increased volumetric flow [56–57]. However, contributions from endothelial dysfunction and arterial stiffness, both of which are well described in SCD, cannot be excluded.

With advancing age, we observed rising SBP and a flattening or declining DBP trend, leading to a widening pulse pressure - a hallmark of arterial stiffening [58–60]. Prior studies have documented higher prevalence of arterial stiffness in SCD compared with controls, attributed to cumulative endothelial damage, impaired NO signalling and cardiovascular remodelling [61–63]. Our longitudinal models demonstrated that while SBP was more volatile and difficult to predict, DBP followed a more defined course, with higher baseline values corresponding to steeper DBP reductions over time. Differences between genotypes may reflect distinct haematocrit-viscosity profiles, where genotypes such as HbSC and HbS/β^+^-thalassaemia, which have higher haematocrit values, are predisposed to higher DBPs due to higher resting cardiovascular resistances.

### Genotype-specific biochemical interactions

Our findings highlighted significant genotype-specific blood pressure responses to circulating levels of renal biomarkers. Notably, HbS/β^0^ and HbS/β^+^ displayed enhanced BP sensitivity to urea and creatinine concentrations, suggesting a more labile vascular role with regards to renal stress. However, the direction of the associations varied, with HbS/β^0^ exhibiting a positive association, whereas HbS/β^+^ had a negative correlation pattern. Creatinine, a surrogate marker of glomerular filtration, is not a direct contributor to BP variability [64–65]. Therefore, elevated creatinine levels serve as a lagging indicator of cardiovascular health and has been linked to increasing hypertension risk in men [66].

In HbSC, we observed blunted responses to most biomarkers despite having higher DBP levels and pervasive HTN-risk, reflecting a dissociation between systemic haemodynamics and microvascular state. However, potassium concentration was inversely and significantly associated with systolic pressure in HbSC, HbS/β^0^ and HbSS. These inverse trends are consistent with the known vasoactive effects of potassium on BP levels via mechanisms involving smooth muscle cell hyperpolarization, endothelium-dependent vasodilation and increased sodium excretion [67–71]. In HbS/β⁺, the discordant response to elevated potassium levels may signal autonomic dysregulation or altered Na⁺/K⁺-ATPase activity leading to a maladaptive pressor response. These patterns are clinically important given the role of potassium levels on vascular haemodynamics, particularly under conditions of volume stress and chronic haemolytic disease.

Urea, the principal nitrogenous waste product generated through hepatic protein catabolism [72], exhibited pronounced genotype-specific effects, particularly pertaining DBP in the HbS/β-thalassaemias. Uremic states contribute to increased vasoconstriction and elevated diastolic pressures through multiple pathways including endothelial dysfunction, oxidant stress and increased sympathetic activity [73–75]. Uremic hypertension and uremic toxins such as para-cresyl sulfate and indoxyl sulfate contribute to increased cardiovascular mortality in chronic kidney disease patients [76], through mechanisms that bear remarkable resemblance to SCD-related vasculopathy.

Collectively, these divergent associations emphasize that similar levels of biochemical derangement may carry different haemodynamic consequences in SCD depending on haemoglobin genotype.

### HTN-risk and comorbid disease

The risk of relative systemic hypertension was coherent across binary and mixed-effects ordered logistic models: cardiac disease (up to 12-fold) and bone pain were associated with high- and severe-risks of HTN, while intercurrent infection was associated with decreasing rHTN-risk. Men retained an approximately 20% greater odds of systemic HTN than age-matched women. Genotype effects persisted wherein, HbSC and HbS/β^+^ exhibited greater odds of rHTN and HTN compared to age and sex-matched individuals with HbSS. These patterns reflect less overt haemolysis and different haematocrit-viscosity relationships between individuals with HbSS and those with clinically milder phenotypes.

Indeed, the cardiac effects were less a surprise than a clinical warning that underlying cardiomyopathy and volume/afterload shifts can constitute increased BP risk in SCD. Once cardiac involvement was present, patients were increasingly likely to occupy the higher BP stratum, even after controlling for age, sex, genotype, biochemical markers and repeated measurements. Individuals with SCD and high or severe rHTN-risk should be considered for proactive volume/afterload management and less conservative BP thresholds than lower-risk individuals.

Acute bone pain amplifies sympathetic tone [77], increases heart rate and transiently increases peripheral resistance [42–43]. Longitudinally, crises correlated with upward BP category shifts. Analgesia-hydration protocols should be paired with short-interval BP evaluation and clear strategies for escalation if BP elevations persist post-crisis. This would enable clinicians to distinguish reflexive BP surges, and transient hypotension, from baseline resting values.

We observed a counterintuitive reduction in BP risk in persons presenting with intercurrent infections. However, two possibilities include: i. cytokine-NO pathway associated systemic vasodilation, and ii. the remedial effects of clinical care as infected patients are likely to receive earlier fluid replacements, receive antipyretics and have regular BP measurements taken with prompt correction of dehydration.

The stepwise age effect post-40 years in this cohort was indicative of arterial stiffening. Men had higher odds which supports established sex differences in vascular tone, RAAS metabolism and cardiometabolic risk.

### Blood pressure variability over time

Our data demonstrated that systolic and diastolic blood pressures behave differently over time. Greater SBP instability compared with the more steady downward trend of DBP, likely reflects the combined influence of arterial stiffness, sympathetic tone, and volume-sensitive mechanisms that differentially affect large-artery versus resistance-artery function in SCD. The negative correlation between DBP intercepts and slopes suggests that patients with initially higher diastolic pressures may undergo compensatory declines, consistent with progressive microvascular remodelling or the result of clinical care. By contrast, SBP slope heterogeneity highlighted the dominant role of inter-individual variation in ventricular–vascular coupling and arterial compliance. These patterns support the notion that SBP may be a more labile marker of haemodynamic stress, while DBP trend may signal longer-term vascular adaptation. Importantly, the downward DBP trends in high- and severe-risk groups, together with the distinct visit-related increase observed in HbSS, reinforce that BP variability is not random but shaped by both clinical risk and genetic background, underscoring heterogeneity in vascular regulation across SCD genotypes.

## CONCLUSION

In this longitudinal SCD-cohort, blood pressure-risk predominated the viscosity-resistance-cardiac axis: patients with cardiac involvement, frequent pain, older age, male sex, and higher-viscosity genotypes, consistently occupied the high HTN-risk categories. Active infections were often associated with transient reductions in BP, presumably due to inducible vasodilatory states and clinical care - and were not a true representation of baseline-risk. Furthermore, DBP, rather than SBP, was a predictor of future genotype- and sex-specific BP heterogeneity. Collectively, these patterns argue for phenotype-aware BP monitoring and earlier intervention, with special attention given to patients with recurrent vaso-occlusive crises, a history of cardiac complications, and haemoglobin genotypes HbSC and HbS/β^+^.

## IMPLICATIONS FOR PRACTICE

Diastolic pressure more closely tracked genotype differences and clinical state and should therefore be prioritised for risk detection, particularly in HbSC and HbS/β^+^. Visit-to-visit systolic pressure, by contrast, was more volatile, providing little prognostic value. BP thresholds should implement sex-specific cut-points and consider biochemistry panels when refining short-term BP management criteria. Widening pulse pressure in older adults is a prompt for earlier arterial stiffness screening and tailored clinical antihypertensive care. While SBP surges can be expected with pain and SCD crises, the overall differences were minimal. Therefore, treatment escalation should not be based solely on contextual BP fluctuations.

## IMPLICATIONS FOR RESEARCH

Investigators may prospectively examine the DBP intercept-slope relationship as an early indicator of SCD vasculopathy. Additionally, the incorporation of ambulatory blood pressure and pulse-wave velocity profiles could clarify involvement of arterial stiffness versus high-input anaemia contributions to SCD blood pressure dysregulation. Furthermore, to determine if rHTN leads to comorbidity or vice versa and to adjust for the effects of time-varying confounding, longitudinal causal inference studies using marginal structural models (MSMs) or inverse probability of treatment weights (IPTWs) should be employed. Finally, the evaluation of catecholamines during SCD steady and crisis states could test the hypotheses surrounding sympathetic surges as a source of SBP volatility during non-routine visits.

## Data Availability

The data has not been hosted by any repositories as we have concerns regarding security since due to the sensitive nature of uploading patient data. However, we are taking steps to engage with platforms where this risk is mitigated.

## APPENDIX

S1 Table 1 Mean SBP and DBP by age group and genotype in females with SCD

S2 Table 2 Mean SBP and DBP by age group and genotype in males with SCD N = number of observations

S3 Table 3 Distribution of routine and non-routine visits across genotypes and hypertension-risk categories

This table summarizes the distribution of patient counts stratified by genotype, hypertension risk category (Lowest, Higher, Severe), and visit type (routine vs non-routine).

